# The COVID in the Context of Pregnancy, Infancy and Parenting (CoCoPIP) Study: protocol for a longitudinal study of parental mental health, social interactions, physical growth, and cognitive development of infants during the pandemic

**DOI:** 10.1101/2021.05.22.21257649

**Authors:** Ezra Aydin, Staci M. Weiss, Kevin A. Glasgow, Jane Barlow, Topun Austin, Mark H. Johnson, Sarah Lloyd-Fox

## Abstract

**Introduction:** While the secondary impact of the COVID pandemic on the psychological wellbeing of pregnant women and parents has become apparent over the past year, the impact of these changes on early social interactions, physical growth and cognitive development of their infants is unknown, as is the way in which a range of COVID related changes have mediated this impact. This study (CoCoPIP) will investigate: i) how parent’s experiences of the social, medical, and financial changes during the pandemic have impacted pre and postnatal parental mental health and parent-infant social interaction; and (ii) the extent to which these COVID-related changes in parental pre and postnatal mental health and social interaction are associated with fetal and infant development.

**Methods and analysis:** The CoCoPIP study is a national online survey initiated in July 2020. This ongoing study (n = 1700 families currently enrolled as of 6^th^ May 2021) involves both quantitative and qualitative data being collected across pregnancy and infancy. It is designed to identify the longitudinal impact of the pandemic from pregnancy to two years of age, with the aim of identifying if stress-associated moderators (i.e., loss of income, COVID-19 illness, access to ante/postnatal support) impact parental mental health, and in turn, infant development. In addition, we aim to document individual differences in social and cognitive development in toddlers who were born during restrictions intended to mitigate COVID-19 spread (e.g., social distancing, national lockdowns).

**Ethics and dissemination:** Ethical approval was given by the University of Cambridge, Psychology Research Ethics Committee (PREC) (PRE.2020.077). Findings will be made available via community engagement, public forums (e.g., social media,) and to national (e.g., NHS England) and local (Cambridge Universities Hospitals NHS Foundation Trust) healthcare partners. Results will be submitted for publication in peer-reviews journals.

**Strengths and Limitations of this study:**

‐ This is a new cohort of families being followed from prenatal to postnatal (up to 18 months) during the COVID-19 pandemic.
‐ The study involves the collection of quantifiable data to identify the short- and long-term influences of the pandemic on key aspects of infant development.
‐ The study also has a range of open-ended questions for qualitative analysis aimed at exploring familial experiences in more detail.
‐ The data is being collected online and is therefore limited to self- and parent-report measures, with no direct assessment of child development and parental mental health.
‐ Although the sample of families being recruited are diverse in their indices of multiple deprivation (IMD) and geographic location, they may not be fully representative of the wider population.

## INTRODUCTION

The COVID-19 pandemic has resulted in an unprecedented challenge to existing medical, social, and economic institutions, akin to prior natural disasters, war or other periods of hardship, with potentially similar long-term implications for the early development and lifelong health of children whose mothers were pregnant or newly delivered during these periods of significant social and economic upheaval [1]. This is due to the fact that the fetal physiological environment and infant caregiving and social environment are now recognised to play a key role in influencing later biological, physical and neurodevelopmental outcomes [2,3]. With regard to the COVID-19 pandemic, the social distancing restrictions and national lockdowns that were put in place to mitigate its transmission, have had a range of secondary consequences impacting the psychological wellbeing of pregnant women and new parents and the postnatal environment that the infant is born into [4–6]. The shifts in socialization, stress and socio-economic position associated with COVID-19 public health guidance may have exacerbated the feelings of vulnerability, health vigilance and isolation associated with the adjustment to parenting. Heightened anxiety and depression were reported during the national lockdown in the United Kingdom [7], with expectant and new mothers experiencing unique physical and psychological stressors [4] as well as constrained access to resources, especially with regard to family and caregiving support. However, little is currently known about the impact of these COVID-related changes on the development of the infant. The current study aims to address this evidence gap by exploring the relationship regarding the family’s reported experiences of these changes in terms of their impact on their pre and postnatal mental health, and interaction with their infant, and the potential subsequent impact of these changes on the infant’s physical, sensory, affective and cognitive development.

### The secondary impact of COVID-19 on pregnant women

The COVID-19 pandemic has been the biggest public health emergency for over a century, necessitating extreme measures at a societal level to mitigate against death and prevent acute health services from being overwhelmed. However, resulting from this have been a number of secondary consequences ((i.e., increased caregiving demands for children and family members; isolation from family and community due to social distancing; job loss; financial hardship and increased interpersonal stressors or relationship violence [8]), having a disproportionate and significant impact on women of childbearing age [4]. In the UK, the impact on this group of women has also been exacerbated by National Health Service (NHS) guidance that was produced in response to the national lockdown restrictions [9–11], in which hospital-based midwifery services placed limitations on partners being present during ultrasound visits and birth. In addition, most community-based services were discontinued, other than antenatal contact and new baby visits, all of which were required to be provided virtually unless otherwise indicated, with all other contacts being assessed and stratified according to vulnerability or clinical need (e.g., maternal mental health).

These changes not only affected the capacity of practitioners to support women during the perinatal period at a time of significantly heightened stress/distress [6] but also resulted in significant regional variations in access to healthcare and advice for expectant mothers across the UK. The changes have been associated with (i) a fourfold increase in stillbirths attributed to lack of preventive antenatal care [12], (ii) birthing partners denied access to the hospital for the birth or asked to leave immediately following the birth, and (iii) limited access to babies admitted to neonatal intensive care. The NHS has also reported a reluctance on the part of parents to attend postnatal GP checks, due to parental attitudes related to COVID-19 infection (Institute of Health Visiting).

### The impact of COVID-19 on parental mental health and parent-infant interaction

Several online surveys conducted during the first national lockdown indicated that there was a significant increase in antenatal anxiety both in terms of pandemic-related pregnancy stress associated with feeling unprepared for birth due to the pandemic, and stress related to fears of perinatal COVID-19 infection, with one large US survey (n = 4451) showing that around 30% of pregnant women experienced both types of stress [13]. Another US survey (n = 2740) that examined wider sources of stress showed that more than half of women reported increased stress about food running out (59.2%, n = 1622), losing a job or household income (63.7%, n = 1745), or loss of childcare (56.3%, n = 1543). More than a third reported increased stress about conflict between household members (37.5%, n = 1028); and 93% (n = 2556) reported increased stress about getting infected with COVID-19 [14].

A number of online cross-sectional surveys found significantly increased rates of anxiety and depression, based on the use of self-report standardised measures (e.g., Edinburgh Postnatal Depression Scale (EPDS); Hospital Anxiety and Depression Scale (HADS)). For example, a cross-sectional survey of 1,987 pregnant women in Canada in April 2020, found substantially elevated anxiety and depression symptoms, compared to similar pre-pandemic pregnancy cohorts; 37% reported clinically relevant symptoms of depression, and 57% reported clinically relevant symptoms of anxiety [15]. A second Canadian study found that a cohort of pregnant women, who were recruited during the COVID-19 pandemic, were twice as likely to present clinically significant levels of depressive and anxiety symptoms compared with a cohort of pregnant women recruited prior to the pandemic [16]. Early evidence in the UK similarly suggests that the impact on the mental health of pregnant women has been significant with heightened anxiety and depression being reported during the national lockdown (levels of mental distress rising from 18.9% (2018-19) to 27.3% in April 2020, one month into the national lockdown) [4].

This is of concern because there is consistent evidence to suggest that anxiety and depression in pregnancy can have a long-term impact on child development. For example, recent systematic reviews found that antenatal anxiety is associated with a range of adverse perinatal outcomes, including for example, premature delivery and low birthweight [17], in addition to a range of negative child outcomes that can persist into late adolescence, including an increased risk of child behaviour problems [18].

### Early pre- and postnatal experiences in the context of COVID-19 and associated societal restrictions

The limited evidence on the impact of the pandemic and lockdowns on postnatal depression, suggest a similar picture, with around half of mothers caring for babies born during 2020 in the UK, reporting feeling down, lonely and worried, with mental health symptoms exacerbated in mothers who travel to work, had a baby born prematurely or were from a lower-income household [19]. One Australian study that examined all online perinatal support forum posts related to COVID-19, from women between January 27 to May 12, 2020, showed that the content was predominantly negative, with around 63% being very or moderately negative. Negative words that were frequently used in the 831 posts included: “worried” (n=165, 19.9%), “risk” (n = 143, 17.2%), “anxiety” (n = 98, 11.8%), “concerns” (n = 74, 8.8%), and “stress” (n = 69, 8.3%) [20].

Anxiety and depression in the postnatal period have been shown to affect the development of the infant because of the impact on the mother’s interactions with her baby. For example, depressed mothers have been shown to be less sensitively attuned to infants, and less affirming and more negating of their experiences with their infant [21]. Babies of depressed mothers can exhibit deficits in their interpersonal functioning, such as less affective sharing, lower rates of interactive behaviour, poorer concentration, increased negative responses with strangers, and reduced secure attachment at 12 and 18 months [22,23]. Children of these women are also 42% more likely to experience depression by age 16 [24].

There is now increasing evidence regarding the impact of the mother’s bond with her unborn baby, including her mental representations of the baby in pregnancy, in terms of an association with their interactions postnatally [25] including the infant’s attachment status [26]. However, there is currently limited evidence regarding the impact of pandemic-related social restrictions on this prenatal relationship. While a number of studies have found that the pandemic has had an impact on bonding with the baby in the postnatal period [27] and on breastfeeding [28], no studies to date have explicitly examined the impact on the parent-child interaction. Furthermore, concerns have been raised regarding infants limited exposure to infant peers or to diverse social partners other than household members or members of the public wearing masks, in terms of the impact on infant looking and communicative bids during social interactions [29]. Whilst ongoing research examines how the pandemic and the above-mentioned points are affecting children aged 8-36 months, there remains a critical lack of study of development during the period of development referred to as the “baby blind-spot” (from pregnancy to 2 years old age) [29,30].

There remain more questions empirical answers at the present time regarding how ‘stay at home’ orders, lack of access to social support from family members and pandemic-specific stressors might have affected expectant parents and those caring for babies during the national lockdowns. Concerns have been raised about women at risk for domestic violence, without suitable shelter and higher incidence of unemployment due to their role in the retail, caregiving and hospitality workforce [31,32]. It is also important to take into consideration other mediating influences on infant development including poverty [33], maternal education [34], marital discord [35], single parenthood and ethnicity [36].

Overall, little is currently known about the impact of COVID-19 guidance and restrictions on the long-term development of the child. To address this gap in our knowledge, this study has two main aims: i) to examine how parent’s experiences of the social, medical, and financial changes during the pandemic have impacted pre and postnatal parental mental health and parent-infant social interaction; and (ii) to investigate the extent to which these COVID related changes in parental pre and postnatal mental health and social interaction are associated with fetal and infant development.

## STUDY DESIGN

The Covid in the Context of Pregnancy, Infancy and Parenting (CoCoPIP) study, is a national online survey being carried out in the UK, which was widely advertised from July 2020, that continues to actively recruit families for participation. The research comprises a mixed-methods study collecting data inclusive of both; a) validated physical and psychological assessments, and b) open-ended questions to allow the participant to elaborate on their experience in their own words. The CoCoPIP Study addresses four key hypotheses (H1-4) (see *Figure 1*).

**Figure 1:**
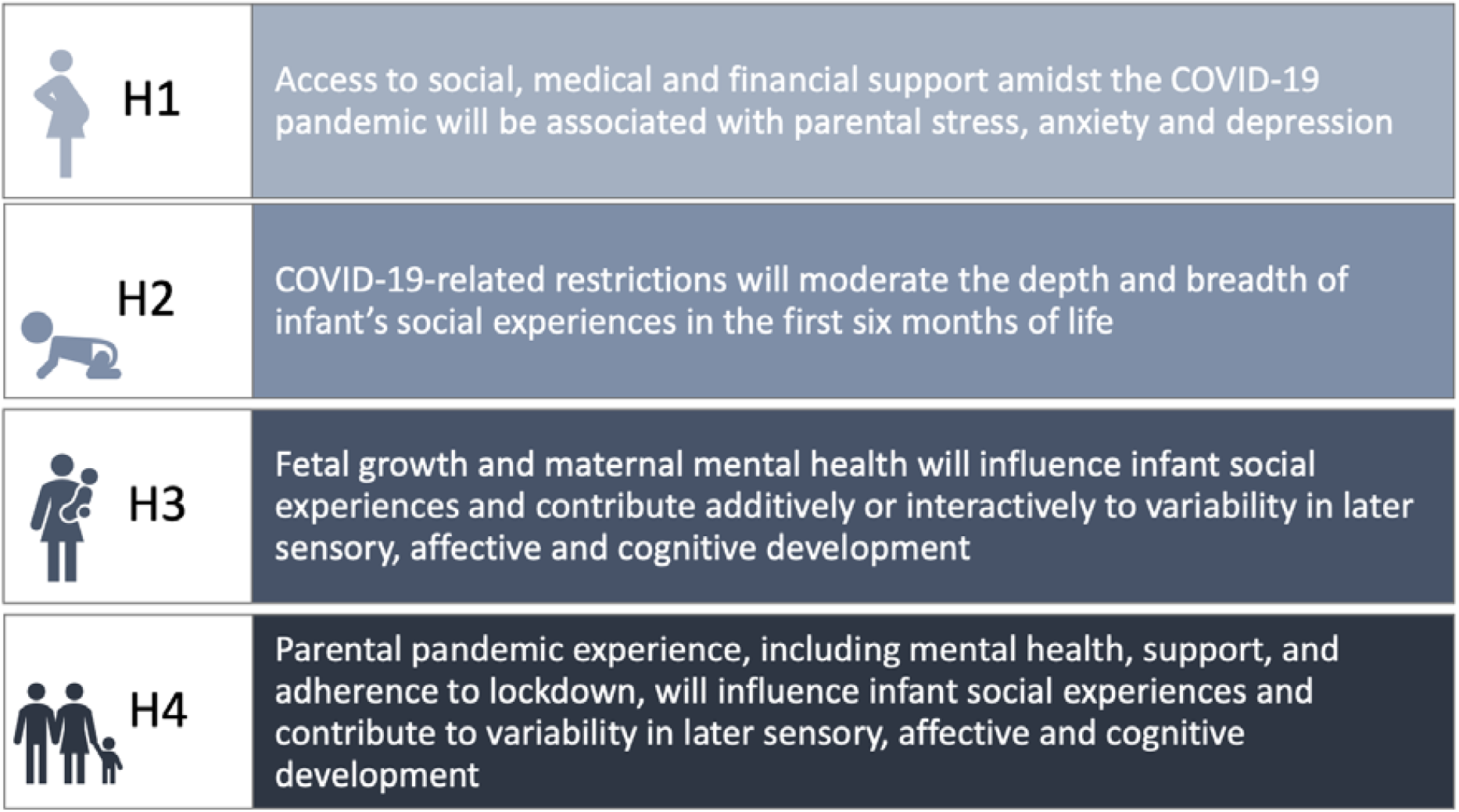
Covid in the Context of Pregnancy, Infancy and Parenting (CoCoPIP) study four key hypotheses.

### Eligibility criteria and recruitment strategies

Eligibility criteria for the study is expectant parents (at any stage of pregnancy) or parents of an infant between the ages of 0-6 months. Either parent can take part, with questionnaires being adapted to the parents’ status as mother or father. The study is open to parents who had a baby shortly before and during the first period of lockdown in the UK (23^rd^ March 2020) as well as continuing to collect information from parents during the current, and future changes in COVID-related health and societal restrictions.

For optimal national representation across the UK, recruitment strategies include (i) targeting NHS antenatal classes and National Childbirth Trust (NCT) groups identified nationally, with an emphasis on areas of low socio-economic status (SES) using the government indices of multiple deprivation (IMD) and rural areas without access to NCT groups, (ii) partnering with NHS/National Institute for Health Research (NIHR) collaborative sites and charity and policy group partners (e.g., The Brazelton Centre UK, Centre for Health and the Public Interest) to widen knowledge of the survey, (iii) posting online via social media platforms (e.g., Twitter) and public sharing to facilitate snowball sampling and (iv) targeting populations experiencing increased local lockdown measures as, and when, COVID-19 rates and related policy change across the UK. Whilst recruitment efforts have been focused on the UK, the survey is currently open to all expectant and new families worldwide. Participation in the survey is incentivized using the offer of a chance to win a £100 digital gift card (on receipt participants are able to select from either an Amazon^®^ or one4all^®^ gift card). A prize is drawn for every 100 participants who complete the survey, giving a 1/100 chance of winning at each time point that they complete.

### Power Calculation

To ensure sufficient power for the study, statistical power calculations (G^*^Power adapted for regression) based on three outcomes, up to three predictors and four co-variables, estimated a minimum sample size for Hypothesis 1 of n = 400 (small effect, f^2^ = 0.02). Statistical power calculations were based on a study of sociodemographic control variables, traumatic event impact scale and pregnancy-specific anxiety [37]. In the same manner, for Hypothesis 2, a minimum sample size of n = 800 (small effect, n = 400 infants x 2 postnatal timepoints) is required, based on ongoing analyses by our group on parent-infant social interaction data [38]. For Hypothesis 3 and 4, we will minimise data loss using post-hoc assignment of families to an accelerated longitudinal design - this requires a minimum sample size of n = 500 (small effect) using a study on acute disasters, parental mental health and infant development [39]. With timing and cross lag accounted for, and attrition rate of 30% - assuming current pattern of 80% of parents’ consenting to be contacted again (as indicated by our pilot survey) – a minimum cohort sample of n = 1500 is required.

### Study Measures

The online survey is logic-dependent and adaptive, only showing questions relevant to the parent’s current situation (e.g., antenatal or with an infant of 2 months of age) in relation to the following six time points: the second and third trimester of pregnancy; infant aged 0-3 months, infant aged 3-6 months; and toddler aged 12- and 18-months. The following data is collected: (i) parental mental health and attitudes, (ii) healthcare access and support during pregnancy and birth, (iii), fetal physical development and infant social and cognitive development, (iv) direct impact of COVID-19 on daily lives and lastly (v) developmental outcomes in infants born during the pandemic. *Table 1* provides an overview of the measures used timepoints and *Table 2* provides a detailed summary of the measures used within the survey.

**Table 1:**
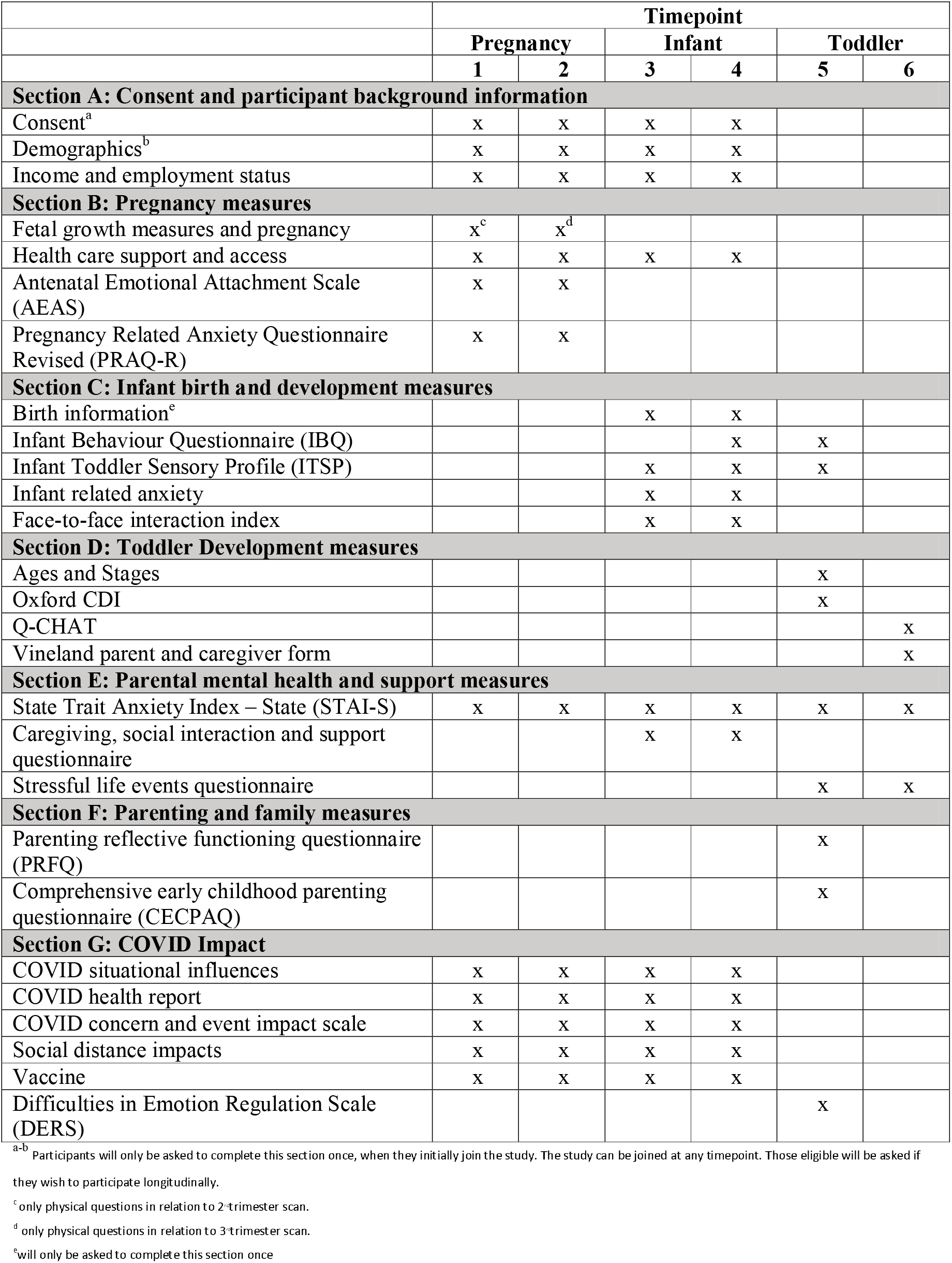
A summary of assessments and questionnaires used separated by time point.

**Table 2:**
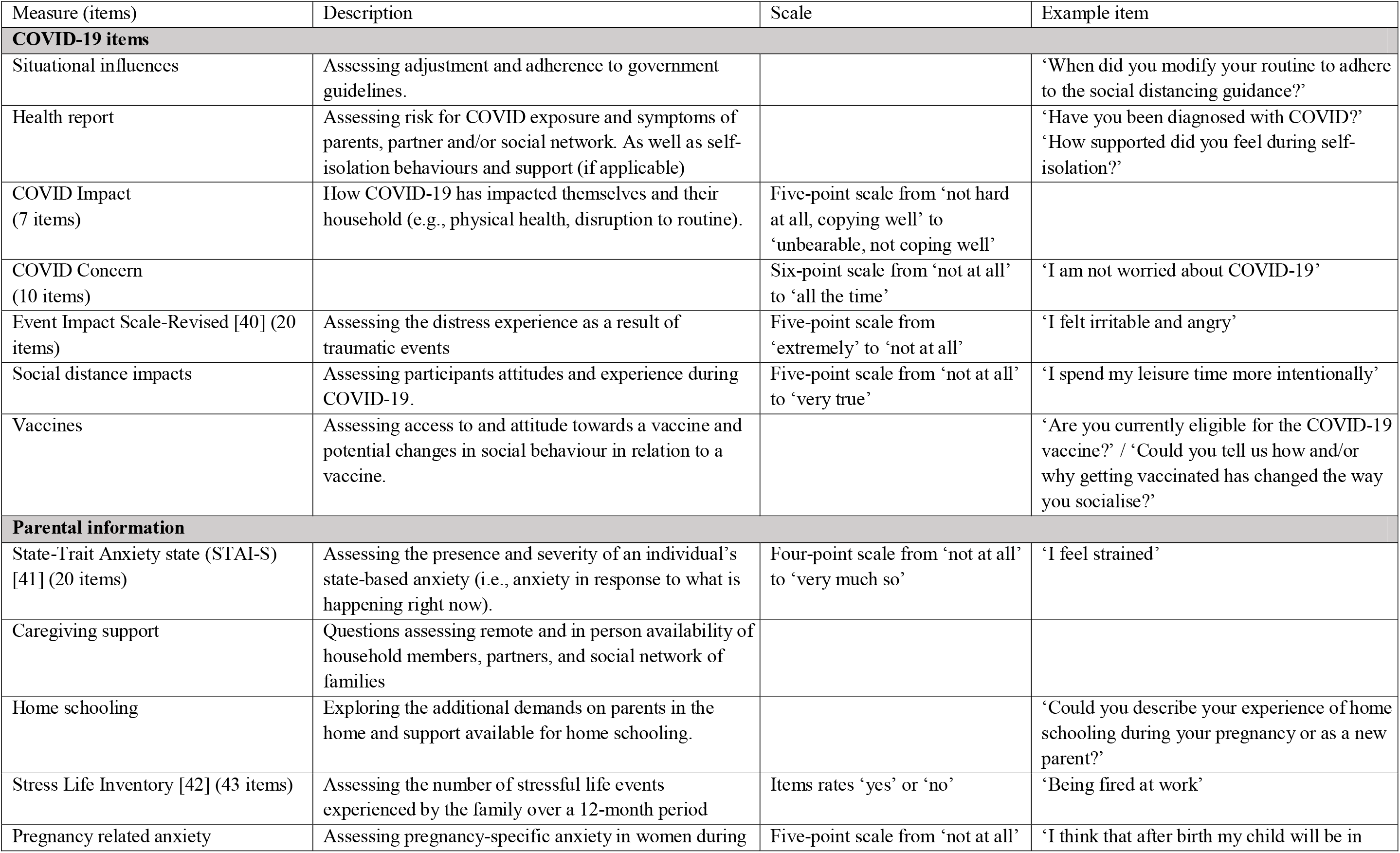
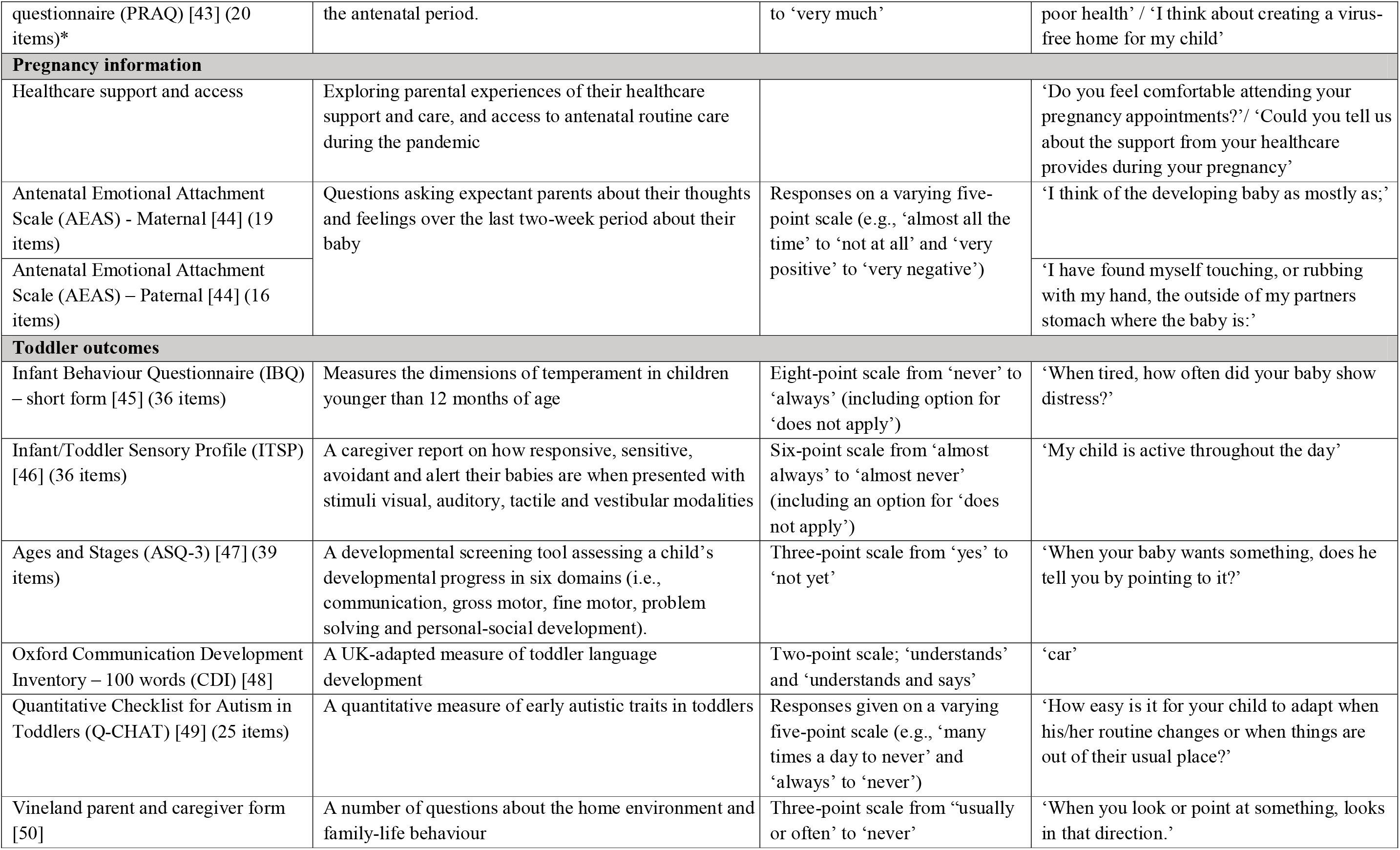
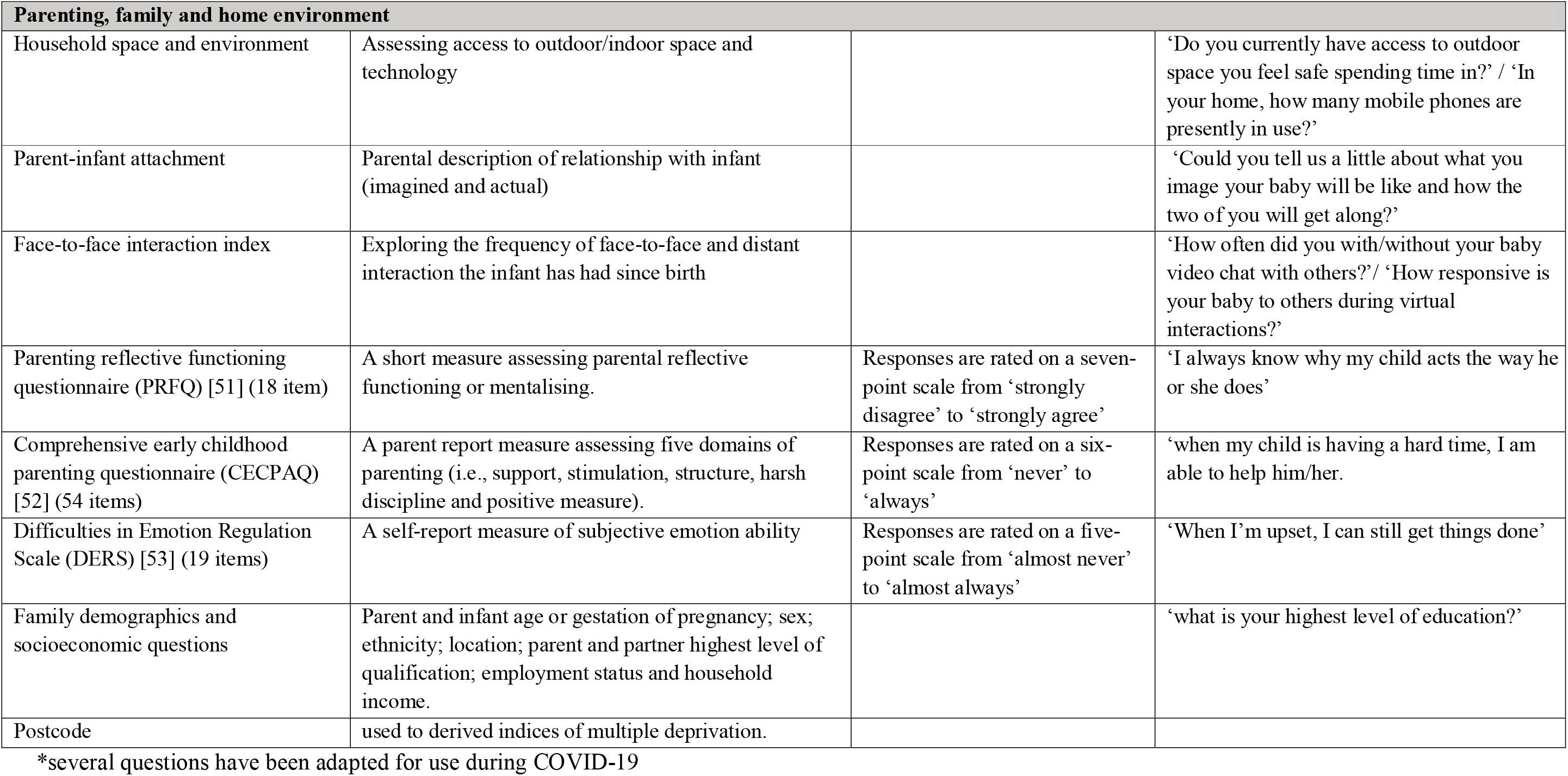
A detailed summary of measures used within the study.

### Follow-ups and reminders

Participants are invited to take part in a follow-up survey at the end of the initial survey. Where they consent to this, they are contacted via email containing a link to the separate online survey. The follow-up survey has been condensed to include follow-up questionnaires only (*see Table 1* and *Figure 2* for participant follow-up flowchart). The appropriate time for follow-up is calculated based on the ages (infant or fetal gestation) provided by the participant at initial recruitment (see *Figure 2* for project timeline).

**Figure 2:**
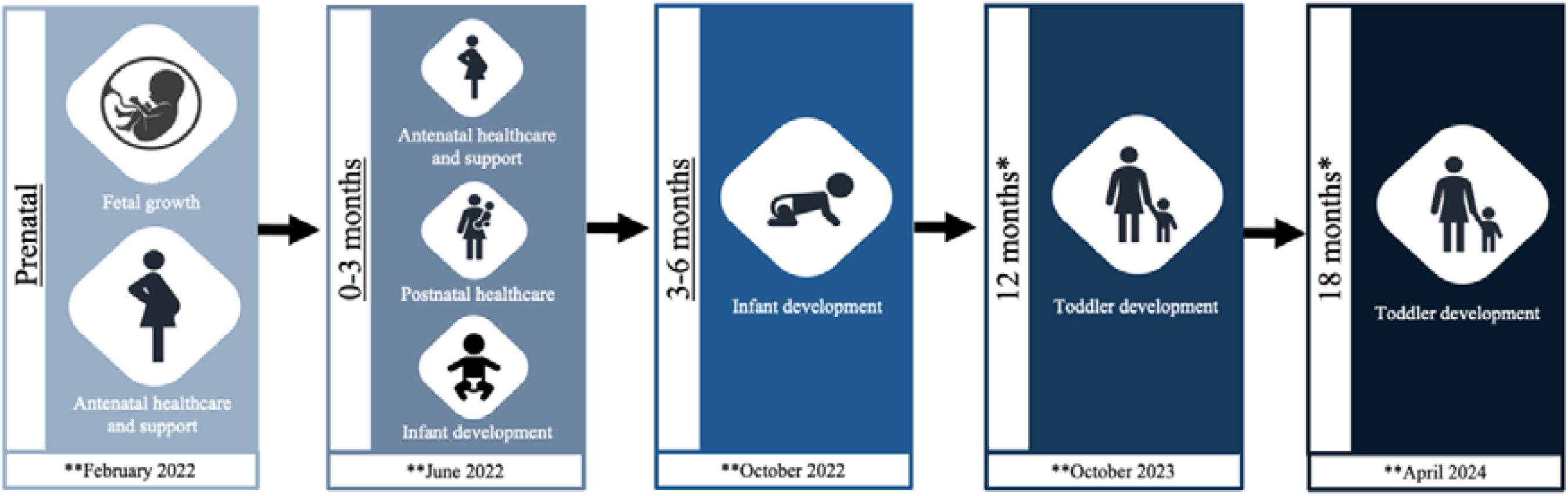
Flow diagram of Covid in the Context of Pregnancy, Infancy and Parenting (CoCoPIP) study follow up participation. A participant can join the study at any of the above 3 underlined timepoints ^*^Participants are only followed up at these two timepoints if they have participated in at least two previous timepoints. ^* *^ Projected participant follow-up completion dates.

## DATA ANALYSIS PLAN

### Quality Control

Ongoing quality control is evaluated bi-weekly. All data is checked for accuracy and invalid data is removed (e.g., one parent responded to the question “what is the gender of your child” as “clownfish” and completed no other question within the survey - this participant was removed from the dataset). Study data is collected and managed using Research Electronic Data Capture (REDCap^®^) tools hosted at the University of Cambridge [54]. REDCap^®^ is a secure, web-based software platform designed to support data capture for research studies, providing 1) an intuitive interface for validated data capture; 2) audit trails for tracking data manipulation and export procedures; 3) automated export procedures for seamless data downloads to common statistical packages; and 4) procedures for data integration and interoperability with external sources. Personal data (e.g., caregiver DOB, email address) is stored securely within a password encrypted electronic databased isolated from the research data. Access to the data is fully audited to ensure data security is governed by a management team and in compliance with ethical guidelines.

### Analysis plan

Overall, we aim to identify which stress-associated moderators (i.e., loss of income, COVID-19 illness, local access to ante/postnatal support) impact significantly on parental mental health, and in turn, infant development.

To address Hypothesis 1 (*see Figure 1*), a combination of quantitative and qualitative analyses will be undertaken. Structural equation modelling (SEM) and hypothesis-driven regressions will explore how multiple aspects of pre-and post-natal family support (social, financial and health) are associated with stress (maternal and paternal) and wider mental health. An inductive approach to data analysis will be undertaken [55] for the open-ended qualitative data, and we will use NVivo (QSR International Pty Ltd) to code the data. Following this, several approaches including thematic [56], sentiment and context content analysis will be undertaken using a natural language processing (NLP) approach [57] (machine learning) to identify forms of social, medical and financial support in relation to the valence of parental attitudes. Regression analyses will then be conducted to understand the directional relationship between resulting latent factors (qualitative responses and quantitative data) and parental mental health.

To address Hypothesis 2, regressions will be used to explore how COVID-19 related restrictions interact with the frequency and forms of social interaction (i.e., face-to-face, digital, at distance) that the infant has, and whether this varies across the first six months of life.

To address Hypothesis 3, regressions will be used to explore the influence of maternal mental health longitudinally on the developing offspring across pre-to post-natal life: from fetal (12-weeks/20 weeks gestational age) to 18 months of age. Standardised z-scores will be created from the collected fetal growth measurements (i.e., head circumference, femur length and abdominal circumference), which will then be transformed into a composite score accounting for gestational age and/or estimated fetal weight at time of scan to be used for analysis. Z-scores will also be computed and used where appropriate within the analysis (e.g., infant-toddler sensory profile).

To address Hypothesis 4, outputs from Hypothesis 1 and 2 (impact of COVID-19 on parental mental health and infant social interactions) will be explored in relation to longitudinal social and cognitive development (e.g., language, motor sensory and the early emergence of developmental conditions) of the infant/toddler across the 0-18 months of life using SEM, full information maximum likelihood to account for missingness, and regression modelling.

## Data Availability

Questionnaires and study goals were made available upon request using the Open Science Foundation platform in July 2020 and made public at https://osf.io/m7zuw/ in August 2020. Qualitative data generated and analysed during the study will not be made publicly available due to ethical and privacy restrictions, however researchers can submit a research proposal to the Data Sharing Management Committee to request access and collaboration. We will either make our datafile associated with each publication available on an opensource platform, following peer review and publication or 12 months after the completion of a follow-up timepoint.

## CURRENT COHORT DESCRIPTION AND DEMOGRAPHICS

Initiated in July 2020, this study is ongoing with n = 1700 families currently enrolled (6^th^ May 2021). Parents can consent to complete the questionnaire up to six times during pregnancy/parenting until their infants are 18 months of age. For those participants who contribute more than one time point (between antenatal and postnatal timepoints ≤ 6 months) an invitation is issued for a follow up to assess their toddler’s development when aged 12 and 18 months (see *Figure 2* for study flow chart).

To date 1700 of families have participated in at least one time-point of the study, with 641 families joining at time points 1-2, 372 families at time-point 3, and 687 families at time-point 4 (see *Figure 2 and Table 1* for time-points). 61% of these families have consented to completing the subsequent follow-up sections of the study. To date 97.4% of respondents identify as mothers, 2.3% as fathers and 0.3% as another parent or caregiver, with the majority of participating families disclosing their ethnicity as white (89.2%). Those participating families who are from the UK have their household information (i.e., household income, location of participating families, index of multiple deprivation and respondent’s education level) described in *Figure 3*.

**Figure 3:**
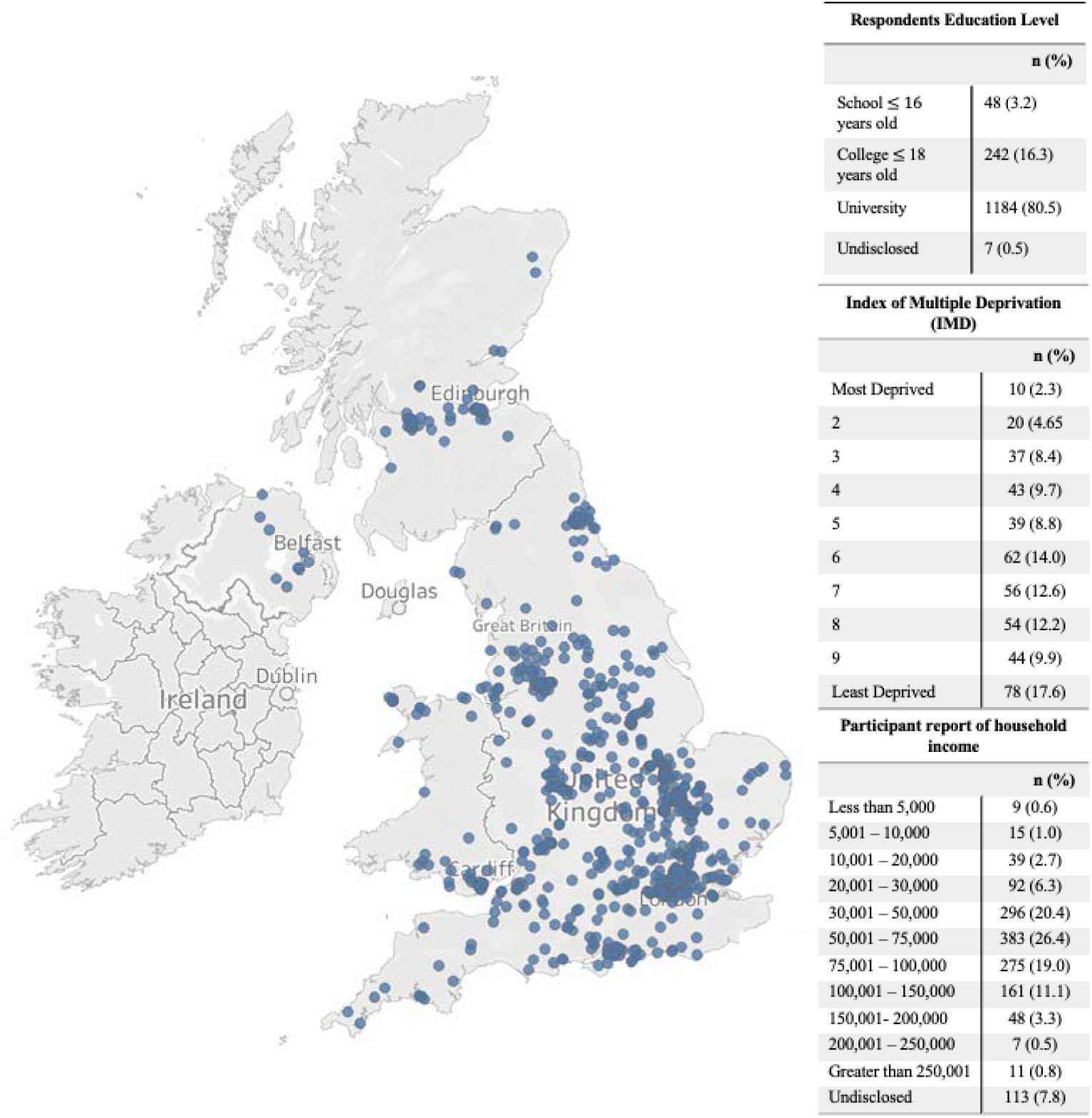
Bubble map depicting spread of participations location in the UK (if postcode was provided) with respondent’s education level, Index of Multiple Deprivation (IMD) and household income breakdown reported on the right.

## ETHICS AND DISSEMINATION

Ethics approval for the survey was given by the University of Cambridge, Psychology Research Ethics Committee (PREC) (PRE.2020.077). All respondents are required to be over the age of 18 years and give electronic informed consent. Caregivers agreeing to be followed-up longitudinally give consent at each timepoint and are made aware that their participation can be stopped at any time within the study. Permissions have been obtained from participants to ensure anonymised data can be made available on open-source platforms.

A proactive dissemination pathway has been established from the outset. We will engage with policy stakeholders (health practitioners/Department of Health) and social media platforms to create discussion around this topic. Dissemination of findings will be via public forums (i.e., social media, media, collaborators family dissemination pathways) and at the national (i.e., NHS England/NHS Improvement, Royal College of Paediatrics and Child Health, Centre for Health and the Public Interest) and local (Cambridge Universities Hospitals NHS Foundation Trust) level. Data will continue to be disseminated throughout the period of the study to promote discussion and raise the profile of the population identified as being one of the most vulnerable and neglected during the pandemic.

To date, qualitative responses from the first five months of data collection have been analysed to explore parents experiences of being pregnant in relation to healthcare access during the pandemic [5]. This was conducted using thematic and sentiment analysis. The initial findings suggest that a range of adverse effects have been experienced by expectant parents in the UK relating to changes in antenatal support and healthcare appointments in response to governmental guidance with regard to social distancing. These findings point to an urgent need to better address the unique health care needs of each pregnant woman going forward.

## Data Sharing Plan

Questionnaires and study goals were made available upon request using the Open Science Foundation platform in July 2020 and made public at https://osf.io/m7zuw/ in August 2020. Study protocol, follow-up questionnaires and statistical analysis code will be uploaded and shared to facilitate data sharing and collaboration, in accordance with Research, Innovation and Science Policy Experts (RISE) EU principles [58]. Data collected will be uploaded to open-source platforms, for utilisation by other researchers and policy stakeholders. In addition, quarterly layman summaries of findings will be reported for parents and the general public on our website and social media platforms. Qualitative data generated and analysed during the study will not be made publicly available due to ethical and privacy restrictions, however researchers can submit a research proposal to the Data Sharing Management Committee to request access and collaboration. We will either make our datafile associated with each publication available on an opensource platform, following peer review and publication or 12 months after the completion of a follow-up timepoint.

## Acknowledgements

We are extremely grateful to all those families who gave their time to participate and to Esther Adememo, Maddie Walton and Zahra Khan who worked on the CoCoPIP study during their undergraduate and master’s studies at Cambridge.

## Author statement: Ezra Aydin

Conceptualization, Methodology, Writing - Original Draft. **Staci M. Weiss:** Conceptualization, Methodology, Writing - Original Draft. **Kevin A. Glasgow:** Methodology, Visualisation, Writing - Review & Editing **Topun Austin:** Methodology, Supervision, Writing - Review & Editing. **Mark H. Johnson:** Supervision, Funding acquisition, Writing - Review & Editing. **Jane Barlow:** Supervision, Writing - Review & Editing. **Sarah Lloyd-Fox:** Conceptualization, Methodology, Supervision, Funding acquisition, Writing - Review & Editing.

